# Sex-differences in Newly Diagnosed Severe Aortic Stenosis in British Columbia (B.C.) between 2012 - 2022

**DOI:** 10.1101/2024.06.11.24308800

**Authors:** Aishwarya Roshan, Jeffrey Yim, Shamikh Lakhani, Jennifer Wang, Aamiya Sidhu, Eric C. Sayre, Karin Humphries, Janarthanan Sathananthan, David Wood, Michael Y.C. Tsang, Darwin F. Yeung, Christina Luong, Parvathy Nair, Kenneth Gin, John Jue, John G. Webb, Teresa S.M. Tsang

## Abstract

**Background:** Despite its high prevalence, little is known about the effect of sex on the management and outcomes of aortic stenosis (AS). We sought to characterize the effect of sex on the clinical evaluation for and provision of aortic valve replacement (AVR), including surgical (SAVR) and transcatheter aortic valve replacement (TAVR), and subsequent morbidity and mortality outcomes.

**Methods:** A comprehensive chart review was conducted on all patients with a first diagnosis of severe aortic stenosis (AS) at Vancouver General and University of British Columbia Hospitals from 2012 to 2022. Exact chi-square and Kruskal-Wallis tests were used to evaluate variables of interest.

**Results:** A total of 1794 studies met inclusion criteria, comprising 782 females (44%) and 1012 males (56%). Females were significantly older than males at the time of first diagnosis (79 versus 75 years, p<0.001). Females were significantly less likely to be evaluated by the TAVR clinic, cardiac surgeon, or to receive aortic valve intervention (p-value≤0.001).

Females were significantly more likely to be rejected for TAVR due to older age (OR 0.23 (0.07, 0.59)), comorbid conditions (OR 0.68 (0.47, 0.97)), and frailty (OR 0.23 (0.07, 0.59)). Females were significantly more likely to be rejected for SAVR on the basis of frailty (OR 0.66 (0.46, 0.94)). Females also had significantly higher rates of 1-year mortality, hospitalization, and heart failure hospitalization compared to males (p-values < 0.05).

**Conclusion:** Our data suggest significant sex-based discrepancies in the management of AS. Females with severe AS are diagnosed later in life and are less likely to be evaluated for valve intervention. They are less likely to receive intervention due to older age, frailty, and multimorbid conditions. Further research is warranted for more effective identification and follow up of aortic stenosis as well as timely referral for AVR, where appropriate, especially of females.

## Introduction

Aortic stenosis (AS) is a common and progressive valvular heart disease worldwide with significant morbidity and mortality.^1^ Over the past several years, the transcatheter aortic valve replacement (TAVR) has gained traction globally as a safe and effective alternative to surgical aortic valve replacement (SAVR) for patients deemed non-optimal candidates for the latter.

TAVR has been shown to provide morbidity and mortality benefits for both women and men.^2–5^ The data on the impact of sex on the practice patterns of AS management and outcomes remains unclear.^6^ Vancouver is one of Canada’s pioneering centres for TAVR innovation and implantation. We sought to characterize the effect of sex on specialist referrals, clinical evaluation for and provision of aortic valve replacement (AVR), including SAVR and TAVR, reasons for non-referral for intervention, as well as outcomes. We sought to understand the impact of sex on the management of AS to facilitate the improvement of service delivery and cardiovascular outcomes.

## Materials and Methods

### Study Design and Selection

The study procedure and protocols were designed in accordance with the Declaration of Helsinki and received approval from the University of British Columbia institutional review board. A retrospective chart review was conducted of all patients with a new diagnosis of severe AS between January 1, 2012, and December 31, 2022, at a tertiary care center. Consecutive transthoracic echocardiograms (echo) meeting criteria were obtained from *Syngo (version 20)*, a secure, password-protected imaging archiving system hosting echocardiograms from both facilities between January 1, 2012, and December 31, 2022.

Clinical data were abstracted from local electronic medical records. Variables of interest were abstracted including age, sex, and the presence or absence of coronary artery disease, dyslipidemia, previous myocardial infarction, previous heart failure, atrial fibrillation, ischemic stroke, hypertension, chronic kidney disease, permanent pacemaker, peripheral vascular disease, chronic obstructive pulmonary disease, and permanent pacemaker. Symptoms at echocardiogram diagnosis were quantified using the New York Heart Association (NYHA) classification scale for heart failure as well as the Canadian Cardiovascular Society (CCS) angina scoring systems. Patients were classified based on whether they were hospitalized during the time of diagnosis or diagnosed as an outpatient.

### Clinical Endpoints

Data were collected on the dates of the initial cardiology visit, TAVR consultation referral, TAVR consultation, surgical consultation referral, surgical consultation, and date of AVR (either TAVR or SAVR). Wait times were calculated, including the time from diagnosis to TAVR/surgical consultation visit, time from diagnosis to AVR, and time from TAVR/SAVR consultation referral to AVR. One-year major adverse cardiovascular outcomes, defined as the development of stroke, persistent atrial fibrillation or flutter, all-cause hospitalization, heart failure hospitalization, or all-cause mortality within one year of the echocardiographic diagnosis of AS, were abstracted from electronic medical records. Reasons for non-referral to both TAVR and surgical consultation, as well as reasons for not receiving TAVR or SAVR, were analyzed and compared between males and females.

### Statistical Analysis

Descriptive statistics were computed for demographic, baseline, and clinical characteristics of patients. Continuous variables were expressed as a median with interquartile range, while categorical variables were expressed as number of events with percentages. Comparisons of demographic, baseline, clinical, and echocardiographic characteristics were made using exact chi-square tests for categorical variables or Kruskal-Wallis tests for continuous data (e.g., time to visit for specialist visits) Analyses were performed using SAS version 9.4 (SAS Institute Inc., Cary, NC, USA), and p<0.05 was considered to be statistically significant.

## Results

Over a 10-year period (2012-2022), 1794 patients [782 female (44%) and 1012 male (56%)] were identified to have severe AS on echo for the first time. Patient characteristics are summarized in Table 1. Females were significantly older than males at time of first diagnosis (79 versus 75, p <0.001). Males were significantly more likely to have been diagnosed with cardiometabolic comorbidities, including dyslipidemia (57.6% versus 48.1%, p-value <0.001), coronary artery disease (55.1% versus 47.6%, p-value 0.002), and diabetes (27.7% versus 23.5%, p-value 0.049), whereas females were significantly more likely to have hypertension (74.2% versus 69.1, p-value 0.020).

**Table 1.**
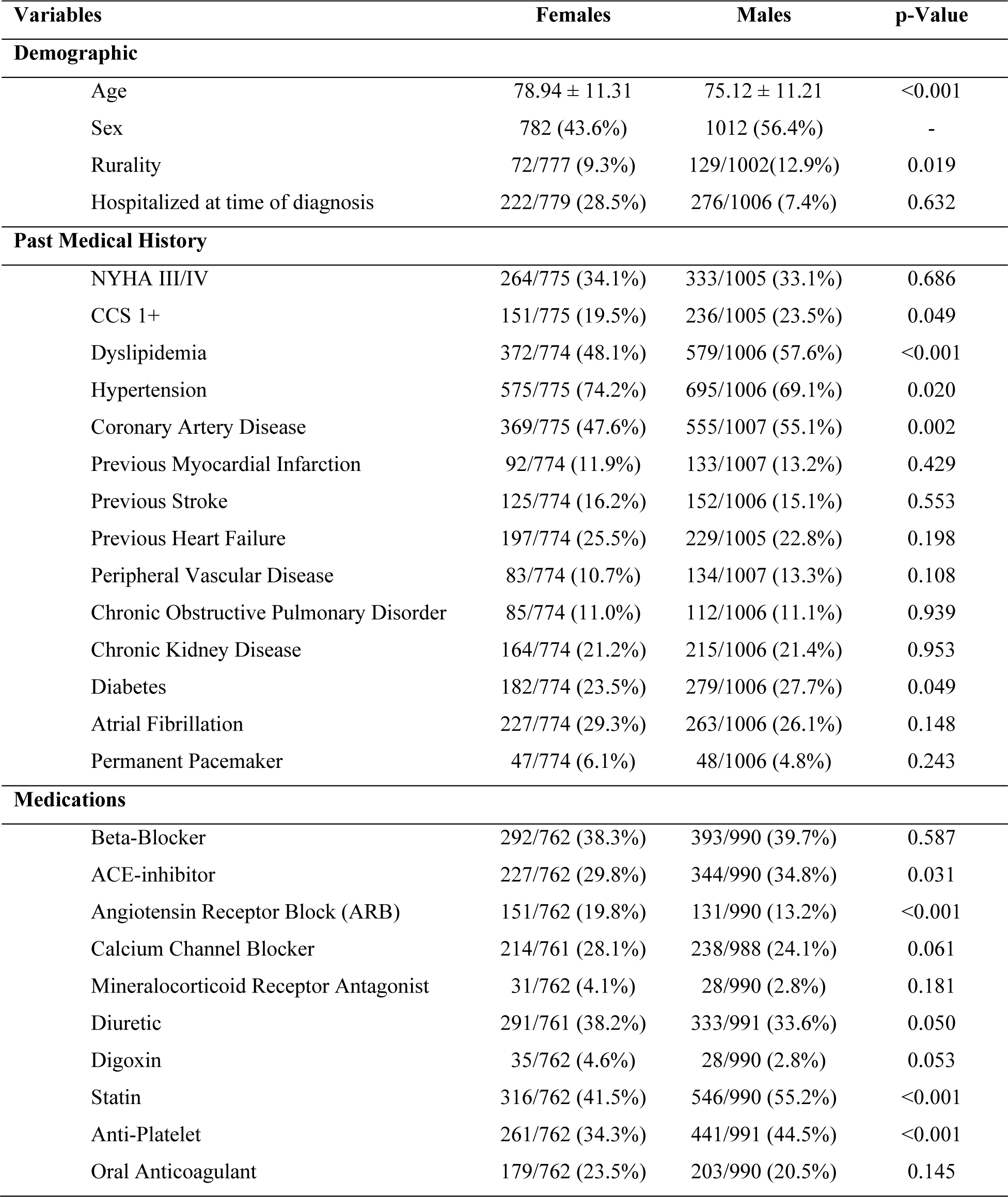
Baseline Characteristics (Demographic, Past Medical History, Medications) of Cohort Stratified by Sex.

Whether a patient was assessed by a specialist (cardiologist, TAVR clinic and/or a surgeon) and/or received intervention (TAVR or SAVR) was tabulated and compared between sexes (Table 2). Males were significant more likely to be evaluated by a surgeon for consideration of SAVR (57.1% versus 47.1%, p-value <0.001), more likely to be evaluated by either a surgeon or the TAVR clinic for consideration of TAVR or SAVR, respectively (75.5% versus 68.3%, p-value 0.001), and more likely to receive an AVR (65.8% versus 52.4%, p-value <0.001). There was no difference in the median wait times between females and males for specialist evaluation and receipt of intervention (Table 3).

**Table 2.**
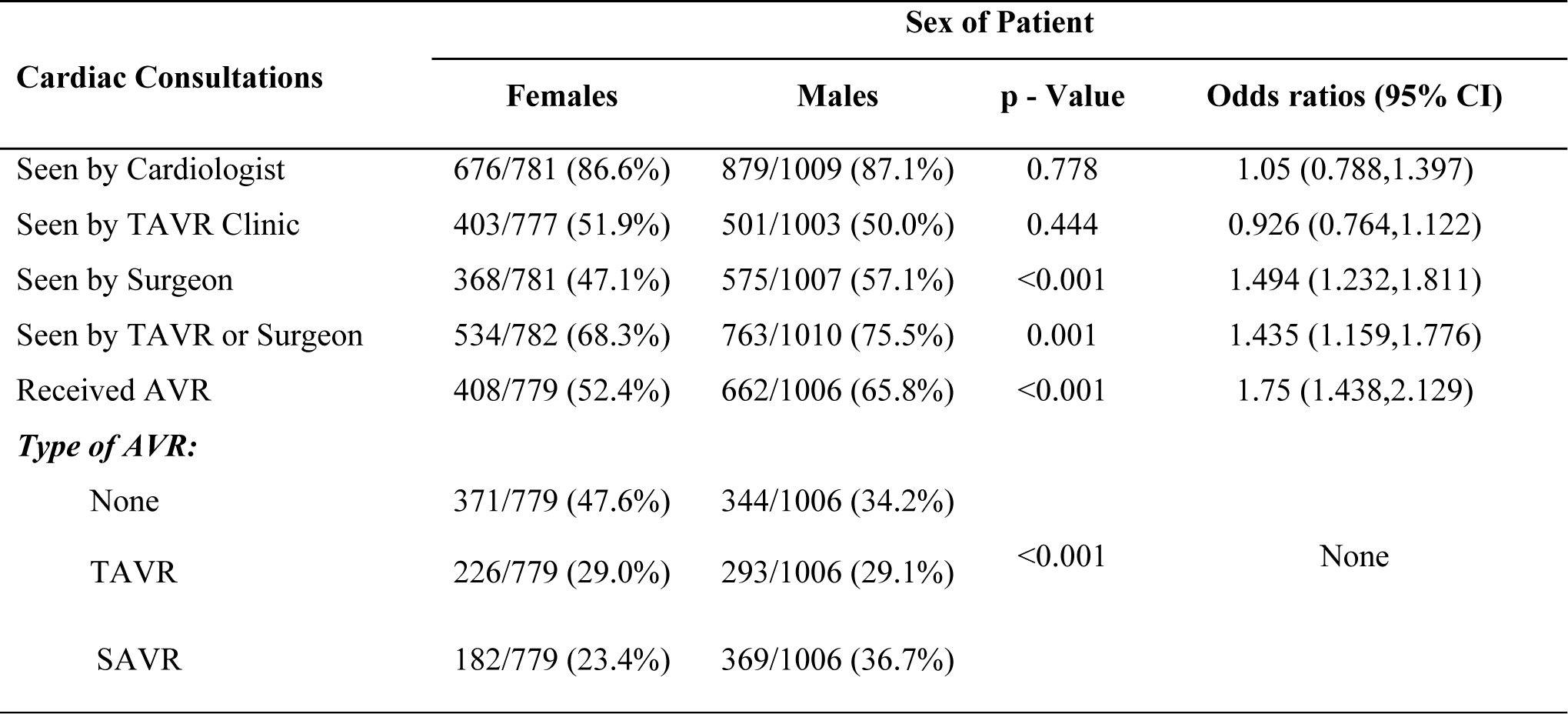
Specialist Assessment and Cardiac Intervention Stratified by Sex.

| Cardiac Consultations | Sex of Patient |  |  |  |
| --- | --- | --- | --- | --- |
|  | Females | Males | p - Value | Odds ratios (95% CI) |
| Seen by Cardiologist | 676/781 (86.6%) | 879/1009 (87.1%) | 0.778 | 1.05 (0.788,1.397) |
| Seen by TAVR Clinic | 403/777 (51.9%) | 501/1003 (50.0%) | 0.444 | 0.926 (0.764,1.122) |
| Seen by Surgeon | 368/781 (47.1%) | 575/1007 (57.1%) | <0.001 | 1.494 (1.232,1.811) |
| Seen by TAVR or Surgeon | 534/782 (68.3%) | 763/1010 (75.5%) | 0.001 | 1.435 (1.159,1.776) |
| Received AVR | 408/779 (52.4%) | 662/1006 (65.8%) | <0.001 | 1.75 (1.438,2.129) |
| **Type of AVR:** |  |  |  |  |
| None | 371/779 (47.6%) | 344/1006 (34.2%) | <0.001 | None |
| TAVR | 226/779 (29.0%) | 293/1006 (29.1%) |  |  |
| SAVR | 182/779 (23.4%) | 369/1006 (36.7%) |  |  |

**Table 3.**
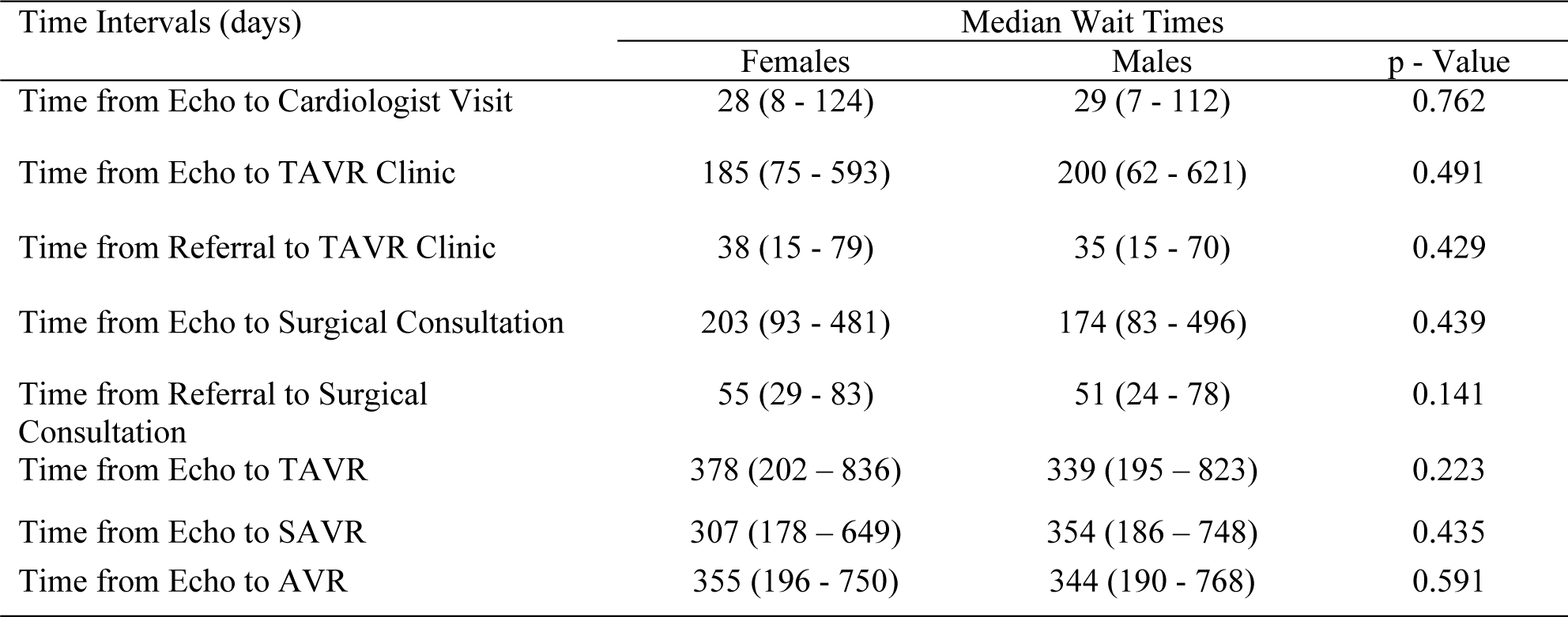
Wait Times Stratified by Sex.

| Time Intervals (days) | Median Wait Times |  |  |
| --- | --- | --- | --- |
|  | Females | Males | p - Value |
| Time from Echo to Cardiologist Visit | 28 (8 - 124) | 29 (7 - 112) | 0.762 |
| Time from Echo to TAVR Clinic | 185 (75 - 593) | 200 (62 - 621) | 0.491 |
| Time from Referral to TAVR Clinic | 38 (15 - 79) | 35 (15 - 70) | 0.429 |
| Time from Echo to Surgical Consultation | 203 (93 - 481) | 174 (83 - 496) | 0.439 |
| Time from Referral to Surgical Consultation | 55 (29 - 83) | 51 (24 - 78) | 0.141 |
| Time from Echo to TAVR | 378 (202 - 836) | 339 (195 - 823) | 0.223 |
| Time from Echo to SAVR | 307 (178 - 649) | 354 (186 - 748) | 0.435 |
| Time from Echo to AVR | 355 (196 - 750) | 344 (190 - 768) | 0.591 |

Along with these differences in specialist care and intervention between the sexes, females were found to have higher rates of 1-year hospitalization and 1-year heart failure hospitalization (Table 4). All findings were highly significant with all p-values <0.015, and most <0.001.

**Table 4.** Cardiovascular Outcomes Stratified by Sex.

| Cardiac Outcomes | Sex of Patient |  |  |  |
| --- | --- | --- | --- | --- |
|  | Females | Males | p - Value | Odds Ratio (95% CI) |
| 1-Year Mortality | 127/779 (16%) | 157/1006 (16%) | 0.696 | 0.949 (0.73, 1.237) |
| 1-Year Hospitalization | 327 (42%) | 359 (36%) | 0.007 | 0.768 (0.631, 0.935) |
| 1-Year HF Hospitalization | 174/777 (22%) | 176/1004 (18%) | 0.012 | 0.737 (0.579, 0.938) |
| 1-Year Persistent Atrial Fibrillation | 59/777 (8%) | 69/1004 (7%) | 0.58 | 0.898 (0.616, 1.312) |
| 1-Year Stroke | 29/778 (4%) | 27/1004 (3%) | 0.221 | 0.714 (0.403, 1.261) |

The reasons for non-referral to specialist evaluation were also examined. While a patient may have more than one reason for not being referred or receiving TAVR and/or SAVR, the largest contributing factor, as delineated by the surgeon/interventionalist on consultation was indicated in our analysis. Females were significantly more likely to be not referred to TAVR clinic for consideration of TAVR due to older age (4% versus 1%, p-value 0.013) and being asymptomatic (12% versus 8%, p-value 0.027). Males were more likely to be not referred to TAVR clinic for requiring concurrent other cardiac surgery (29% versus 24%, p-value <0.001) (Table 5). There were no significant differences between males and females in non-referral to SAVR clinic (Table 6).

**Table 5.** Reasons for Non-Referral to TAVR Clinic.

| Reason | Frequency (n) | Female | Male | p-value |
| --- | --- | --- | --- | --- |
| Age | 20 | 14/376 (4%) | 6/509 (1%) | 0.013 |
| Multiple comorbid conditions | 92 | 47/376 (13%) | 45/509 (9%) | 0.094 |
| Frailty | 33 | 19/376 (5%) | 14/509 (3%) | 0.105 |
| Limited life expectancy | 23 | 12/376 (3%) | 11/509 (2%) | 0.395 |
| Patient decision | 96 | 43/376 (11%) | 53/509 (10%) | 0.662 |
| Concurrent cardiac surgery required | 191 | 45/376 (24%) | 146/509 (29%) | <0.001 |
| Other | 46 | 21/376 (6%) | 25/509 (5%) | 0.760 |
| Low STS score | 168 | 67/376 (18%) | 101/509 (20%) | 0.488 |
| Poor TAVR anatomy | 2 | 2/376 (1%) | 0/509 (0%) | 0.180 |
| Asymptomatic | 83 | 45/376 (12%) | 38/509 (8%) | 0.027 |
| Cognitive dysfunction | 27 | 15/376 (4%) | 12/509 (2%) | 0.172 |
| Patient passed prior | 42 | 16/376 (4%) | 26/509 (5%) | 0.633 |
| Multi-valvular disease | 23 | 15/376 (4%) | 8/509 (2%) | 0.032 |
| Poor functional status | 10 | 5/376 (1%) | 5/509 (1%) | 0.751 |
| Lost to follow up | 29 | 10/376 (3%) | 19/509 (4%) | 0.447 |

**Table 6.** Reasons for Non-Referral to Surgical Evaluation.

| Reason | Frequency (n) | Female | Male | p-value |
| --- | --- | --- | --- | --- |
| Age | 67 | 34/413 (8%) | 33/432 (8%) | 0.800 |
| Multiple comorbid conditions | 235 | 110/413 (27%) | 125/432 (29%) | 0.490 |
| Frailty | 97 | 54/413 (13%) | 43/432 (10%) | 0.132 |
| Limited life expectancy | 26 | 13/413 (3%) | 13/432 (3%) | 1.000 |
| Patient decision | 120 | 57/413 (14%) | 63/432 (15%) | 0.768 |
| Other | 46 | 21/413 (5%) | 25/432 (6%) | 0.762 |
| Poor surgical anatomy | 13 | 8/413 (2%) | 5/432 (1%) | 0.411 |
| Asymptomatic | 85 | 45/413 (11%) | 40/432 (9%) | 0.493 |
| Cognitive dysfunction | 34 | 17/413 (4%) | 17/432 (4%) | 1.000 |
| Patient passed prior | 39 | 15/413 (4%) | 24/432 (6%) | 0.194 |
| Multi Valve disease | 1 | 0/413 (0%) | 1/432 (0%) | 1.000 |
| Too high risk for surgery | 16 | 10/413 (2%) | 6/432 (1%) | 0.319 |
| Poor functional status | 12 | 6/413 (2%) | 6/432 (1%) | 1.000 |
| Lost to follow up | 28 | 10/413 (2%) | 18/432 (4%) | 0.181 |
| Unclear | 26 | 13/413 (3%) | 13/432 (3%) | 1.000 |

Females were significantly more likely to be rejected for TAVR on the basis of older age (4% versus 1%, p-value=0.001), multiple comorbid conditions (13% versus 9%, p-value=0.031), frailty (5% versus 2%, p-value=0.004), having poor TAVR anatomy, and having multivalvular heart disease (5% versus 2%, p-value 0.004). Males, on the other hand, were rejected for TAVR on the basis of requiring concurrent other cardiac surgery (30% versus 15%, p-value<0.001), as well as having a low Society of Thoracic Surgeons (STS) score (21% versus 17%, p-value=0.063) (Table 7).

**Table 7.** Reasons for Not Receiving TAVR.

| Reason | Frequency (n) | Female | Male | p-value |
| --- | --- | --- | --- | --- |
| Age | 26 | 20/555 (4%) | 6/712 (1%) | 0.001 |
| Multiple comorbid conditions | 141 | 74/555 (13%) | 67/712 (9%) | 0.031 |
| Frailty | 46 | 30/555 (5%) | 16/712 (2%) | 0.004 |
| Limited life expectancy | 31 | 16/555 (3%) | 15/712 (2%) | 0.464 |
| Patient decision | 120 | 55/555 (10%) | 65/712 (9%) | 0.699 |
| Concurrent cardiac surgery required | 298 | 83/555 (15%) | 215/712 (30%) | <0.001 |
| Other | 48 | 22/555 (4%) | 26/712 (4%) | 0.882 |
| Low STS score | 247 | 95/555 (17%) | 152/712 (21%) | 0.063 |
| Poor TAVR anatomy | 32 | 21/555 (4%) | 11/712 (2%) | 0.032 |
| Asymptomatic | 93 | 49/555 (9%) | 44/712 (6%) | 0.082 |
| Cognitive dysfunction | 31 | 17/555 (3%) | 14/712 (2%) | 0.271 |
| Patient passed prior | 71 | 31/555 (6%) | 40/712 (6%) | 1.000 |
| Multi Valve disease | 37 | 25/555 (5%) | 12/712 (2%) | 0.004 |
| Poor functional status | 15 | 7/555 (1%) | 8/712 (1%) | 1.000 |
| Lost to follow up | 31 | 10/555 (2%) | 21/712 (3%) | 0.205 |

Females were also significantly more likely to be rejected for SAVR on the basis of frailty (15% versus 10%, p-value=0.019) (Table 8).

**Table 8.** Reasons for Not Receiving SAVR.

| Reason | Frequency (n) | Female | Male | p-value |
| --- | --- | --- | --- | --- |
| Age | 91 | 44/594 (7%) | 47/639 (7%) | 1.000 |
| Multiple comorbid conditions | 377 | 179/594 (30%) | 198/639 (31%) | 0.757 |
| Frailty | 152 | 87/594 (15%) | 65/639 (10%) | 0.019 |
| Limited life expectancy | 30 | 14/594 (2%) | 16/639 (3%) | 1.000 |
| Patient decision not to proceed | 153 | 72/594 (12%) | 81/639 (13%) | 0.796 |
| Other | 47 | 21/594 (4%) | 26/639 (4%) | 0.658 |
| Poor surgical anatomy | 42 | 22/594 (4%) | 20/639 (3%) | 0.639 |
| Asymptomatic | 91 | 48/594 (8%) | 43/639 (7%) | 0.385 |
| Cognitive dysfunction | 43 | 21/594 (4%) | 22/639 (3%) | 1.000 |
| Patient passed prior | 47 | 17/594 (3%) | 30/639 (5%) | 0.103 |
| Multi Valve disease | 1 | 0/594 (0%) | 1/639 (0%) | 1.000 |
| Too high risk for surgery | 64 | 28/594 (5%) | 36/639 (6%) | 0.521 |
| Poor functional status | 33 | 16/594 (3%) | 17/639 (3%) | 1.000 |
| Lost to follow up | 31 | 10/594 (2%) | 21/639 (3%) | 0.100 |
| Unclear | 31 | 15/594 (3%) | 16/639 (3%) | 1.000 |

A multivariable analysis was conducted to evaluate independent predictors of receiving TAVR. Amongst those receiving AVR, females were more likely to receive TAVR compared to their male counterparts [OR of 1.47 (1.07,2.01); p=0.016] in the multivariate analysis.

## Discussion

Our data suggests that significant sex differences existed across our large cohort in the management of severe AS. Specifically, males had a higher comorbidity burden compared to females. They were more likely to have dyslipidemia, coronary artery disease, and diabetes, as well as require a statin and anti-platelet agent. This is consistent with prior research that suggests males have more cardiovascular comorbidities including myocardial infarction, stable angina, ischemic stroke, peripheral arterial disease, heart failure, and cardiac arrest.^7,8^

However, despite the higher burden of cardiovascular disease amongst males, the literature points to females having higher rates of morbidity and mortality post-acute coronary syndrome diagnosis, particularly within ST elevation myocardial infarctions.^9–11^ In our study, females were more likely to require hospitalization for heart failure and for other reasons, when compared to males within 1 year of their AS diagnosis.

The diagnosis and treatment for female patients at a more advanced age has long been observed in the context of acute coronary syndrome (ACS) and is a leading explanation for the mortality differences observed between females and males.^10^ Females are less likely to undergo diagnostic procedures, such as coronary angiography; less likely to receive treatments, such as percutaneous coronary intervention, coronary artery bypass graft, or optimal medical therapy; and more likely to have prehospital delays in obtaining adequate care for their ACS.^10–16^ Likewise, our study showed that females were less likely to be evaluated by the TAVR clinic or a surgeon, and less likely to receive an AVR, which may explain the differences in morbidity between the sexes.

Females were significantly less likely to be referred for and/or receive TAVR due to old age. This held the strongest statistical significance out of all reasons. While echocardiographic parameters were not directly analyzed in our paper, our finding of reduced female patient referral coupled with prior research delineating differences in LV structure, function, and hemodynamics between the sexes underlines the importance of developing sex-specific criteria in defining AS severity.^17^ The next most important reason for non-intervention in females was frailty. Even amongst individuals who received surgical assessment, females were significantly more likely to be rejected for SAVR on the basis of frailty. Both SAVR and TAVR confer significant mortality and morbidity benefits, and thus, understanding the reasons for lack of treatment is pivotal in enabling physicians to provide optimal care. Both age and frailty are implicated in the lack of intervention in female patients. One of the highest predictors of frailty is advanced age.^18^ In our cohort, females are diagnosed with severe AS at a later age, when compared to males. It is unclear if they developed symptoms earlier, as well. In light of these findings, consideration of sex-based triaging criteria for intervention for improvement of outcomes could be considered.

One of the most significant reasons for not pursuing SAVR within our cohort was patient preference, which is also identified within the literature amongst female patients.^19–22^

Despite being a large study, limitations of our study include that it was conducted within a single institution with a single electronic medical record. This project therefore does not account for hospitalizations, diagnoses, and events obtained outside of the Vancouver Coastal Health jurisdiction and assumed that patients of our study only obtained care within this health authority.

### Conclusion

There appeared to be significant sex-based discrepancies in the management of AS. Females were diagnosed with severe AS later in life. They were less likely to be evaluated for valve replacement (both TAVR and SAVR) and upon evaluation, they were also less likely to receive this intervention, due to their older age, frailty, and multimorbid conditions. Greater efforts are needed to identify females with severe AS earlier in disease progression. Further research is required to determine whether the lack of referral and receipt of AVR amongst females are independent predictors of their significantly higher morbidity and mortality outcomes, and whether triaging criteria for intervention incorporating sex will improve outcomes for female patients.

## Data Availability

The data that support the findings of this study are available from the corresponding author upon reasonable request.

## Acknowledgments

The corresponding author affirms that everyone listed has contributed significantly to this work. The authors had access to all the study data, take responsibility for the accuracy of the analysis, and had authority over this article. The corresponding author confirms that all authors read and approve the article.

## Sources of Funding

None

## Disclosures

None

## Notes

### Competing Interest Statement

The authors have declared no competing interest.

### Funding Statement

No funding received.

### Author Declarations

University of British Columbia Research Ethics

